# Metacognitive insight into cognitive performance in pre and early-stage Huntington’s disease

**DOI:** 10.1101/2021.10.25.21265369

**Authors:** Samuel RC Hewitt, Alice J White, Sarah L Mason, Roger A Barker

## Abstract

**Objectives:** Insight is an important predictor of quality of life in Huntington’s disease and other neurodegenerative conditions. However, estimating insight with traditional methods such as questionnaires is challenging and subject to limitations. This cross-sectional study experimentally quantified metacognitive insight into cognitive performance in Huntington’s disease gene-carriers.

**Methods:** We dissociated perceptual decision-making performance and metacognitive insight into performance in healthy controls (n=29), premanifest (n=19) and early-manifest (n=10) Huntington’s disease gene-carriers. Insight was operationalised as the degree to which a participant’s confidence in their performance was informative of their actual performance (metacognitive efficiency) and estimated using a computational model (HMeta-d).

**Results:** We found that pre and early-manifest Huntington’s disease gene-carriers were impaired in making perceptual decisions compared to controls. Gene-carriers required more evidence in favour of the correct choice to achieve similar performance and perceptual impairments were increased in those with manifest disease. Surprisingly, despite marked perceptual impairments, Huntington’s disease gene-carriers retained metacognitive insight into their perceptual performance. This was the case after controlling for confounding variables and regardless of disease stage.

**Conclusion:** We report for the first time a dissociation between impaired cognition and intact metacognition (trial-by-trial insight) in the early-stages of a neurodegenerative disease. This unexpected finding contrasts with the prevailing assumption that cognitive deficits are associated with impaired insight. Future studies should investigate how intact metacognitive insight could be used by some early Huntington’s disease gene-carriers to positively impact their quality of life.

## INTRODUCTION

Huntington’s disease (HD) is a neurodegenerative disorder caused by a CAG expansion in exon 1 of the Huntingtin gene (1). HD gene-carriers are currently diagnosed with manifest disease when abnormal movements emerge, but true disease onset begins years earlier. The cognitive features of HD develop in the premanifest stage and include impaired executive cognition (planning, reasoning, working memory and attention (2)), psychomotor processing speed, visuospatial functions and emotion recognition (3). Patients tend to perceive their abilities differently from their carers; typically underestimating their impairments when asked to explicitly reflect on them (4). We refer to this as global insight, and it is thought that HD patients become increasingly impaired as disease burden increases.

However, studies of global metacognitive insight such as those which rely on self-report are subject to several confounding influences which limit their interpretability. This is because global insight is a complex concept which is influenced by many individual differences. For instance, systematic response biases (e.g., optimism), personality dimensions or temporary psychological states (e.g., trait-anxiety or stress) and other critical cognitive functions (e.g., episodic memory) can all affect the way that patients report on themselves. Here, we specify metacognitive insight as the accuracy of reflection on performance in a cognitive task (i.e., insight into task performance on a trial-by-trial basis). This has been referred to as local metacognition and is distinct from global insight (5). Global insight is hierarchically more abstract,, spans longer timescales and captures how we feel about performance broadly, for example, across an entire task, a cognitive domain or in daily life (6).

Local metacognitive insight has been associated with neural substrates which are also affected early on in HD. For example, in healthy controls it has been associated with increased anterior and medial prefrontal cortex activity (6–8) and altered hippocampal myelination (9). Premanifest HD gene-carriers are known to exhibit grey matter loss in the prefrontal cortex (10) and hippocampal dysfunction is reported with late premanifest and manifest HD (11). However, local metacognitive insight, as defined here, has not been explicitly tested in HD.

In this study, we dissociated perceptual cognitive performance from metacognitive insight into performance, in premanifest and early-manifest HD, and age- and sex-matched healthy controls. In order to quantify metacognitive insight independently of confounding influences, we controlled perceptual decision-making performance across participants and employed an established computational model of metacognition (12). We hypothesised that HD gene-carriers would show impairments in decision-making performance, and this would be compounded by a reduction in metacognitive insight into performance. We predicted that these impairments would be significantly greater in those with early-manifest disease.

## METHODS

### Participants

Sixty-three participants completed this study: 14 patients with early-manifest HD, 20 premanifest gene-carriers and 29 healthy controls between September 2019 and November 2020. All HD gene-carriers were genetically confirmed (CAG ≥ 36). Patients were defined as having early-manifest disease when they had a Unified Huntington’s Disease Rating Scale (UHDRS) total motor score > 5 (13). The groups were matched for age and sex. Inclusion criteria were Mini Mental State Examination (MMSE) score > 26 (normal range) and UHDRS initiation and saccade velocity total scores less than or equal to 1 (indicating minimal impairment in one domain only; maximum score is 16). Therefore, all included participants with gene-positive HD had no global cognitive or saccadic impairments as detected during examination by an experienced Consultant Neurologist (R.A.B). Exclusion criteria were any significant comorbid psychiatric or neurological diagnoses. Participants with HD were recruited from the HD Clinic at the University of Cambridge and Cambridge Universities Hospitals NHS (National Health Service) Foundation Trust. Controls were recruited from the local community. All participants gave prior written informed consent. This study was approved by the Oxford South Central-C Research Ethics Committee and the Medical Health Research Authority in the United Kingdom. Clinical data were collected and managed using REDCap (Research Electronic Data Capture) electronic data capture tools hosted at the University of Cambridge (14,15). This study is reported in accordance with the STROBE reporting checklist (16).

### Stimuli and Procedure

We employed a task previously used to separately assess perceptual decision-making and metacognition (7), implemented in MATLAB using Psychtoolbox (17). The code used to run the task is available online (12). Participants were required to make an alternative forced-choice judgement about which of two briefly presented (0.7secs) circles contained more dots, for which there was no response time limit. One of the two circles contained 50 dots while the other circle contained a number bounded between 1 and 100. On each trial, this was followed by a confidence rating which had to be made within 4s of the confidence scale being shown. All stimuli were high contrast (white on black; Figure 1). A one-up two-down staircase procedure equated performance across participants based on response accuracy by manipulating the stimulus strength (Δ dots) such that performance was constant (∼71%, Figure 2A). The staircase procedure was initiated during a practice phase which provided feedback on decision accuracy.

**Figure 1.**
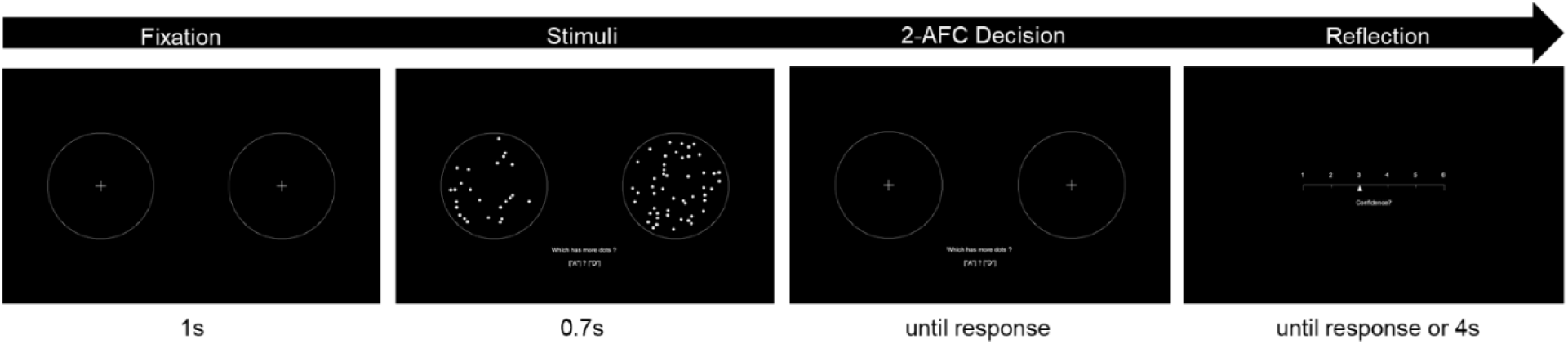
Meta-dots task. Participants are required to make an alternative forced choice judgement (2-AFC) about which of the two stimuli(circles) contain more dots. This is immediately followed by a confidence rating. Figure adapted from Fleming et. Al., 2014.

**Figure 2.**
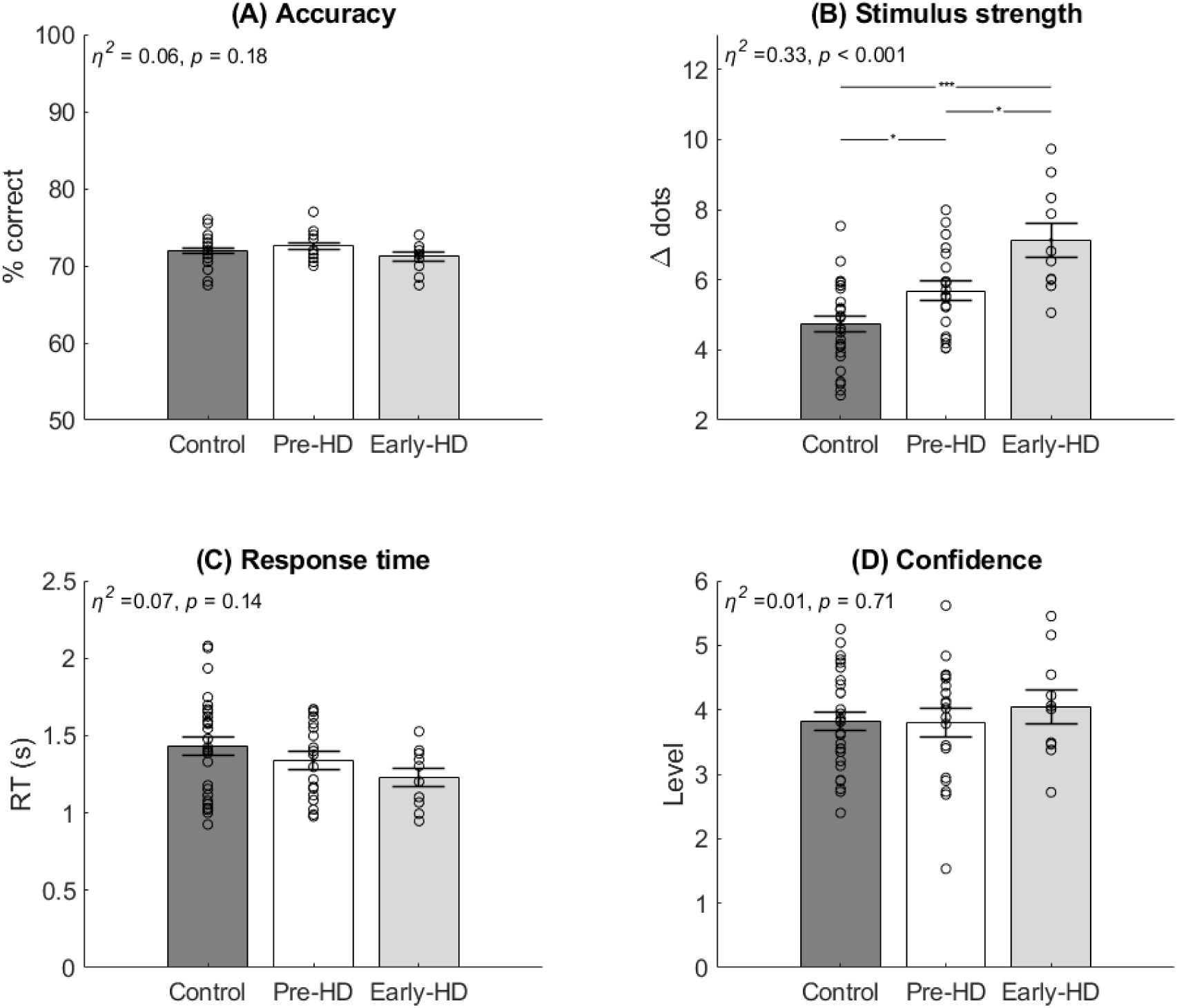
Behavioural data. (A) Accuracy was controlled across the groups at approximately 71%. (B) Stimulus strength (Δ dots) was significantly increased in the early-manifest group, compared with both groups, and also in the premanifest group compared to the control group. (C) No significant difference in mean response time. (D) No significant difference in mean confidence. Bars = mean ± SEM (errors). Circles = individual mean values. *Bonferroni corrected *p* <0.05. ***Bonferroni corrected *p* <0.001. *η*^*2*^ = ETA squared effect size. Abbreviations: pre-HD = premanifest Huntington’s disease, early-HD= early manifest Huntington’s disease.

Feedback was not given after the practice. The experiment was divided into 8 blocks of 25 trials, separated by a break of length determined by the participant.

Participants also completed the Hospital Anxiety and Depression Scale (HADS), Mini Mental State Examination (MMSE) and the National Adult Reading Test (NART), which was used to calculate predicted verbal IQ.

### Metacognitive insight

We used metacognitive efficiency (M-ratio) as an index for metacognitive insight across premanifest HD, early manifest-HD, and healthy controls. M-ratio is an established marker of metacognition based on signal detection theory (16,17). M-ratio describes how much of the available signal (i.e., a participant’s perceptual sensitivity, *d’*) is captured by their confidence about their performance on that trial. Specifically, M-ratio is the ratio between metacognitive sensitivity (*meta-d’*) and perceptual sensitivity (*d’*). As such, this method controls for differences in perceptual ability as well as response biases (e.g., repeatedly high confidence) and is well-suited to compare metacognitive insight in clinical groups. An M-ratio of 1 would represent optimal sensitivity to perceptual performance. If M-ratio < 1, there is some noise in the confidence ratings, such that the individual does not exploit all the available perceptual signal for their metacognitive judgement. If M-ratio > 1, this implies that the individual can draw on additional information about themselves or the task (beyond the available perceptual signal) (20). We estimated M-ratio using a hierarchical modelling approach implemented in an openly available MATLAB toolbox (HMeta-d, (18)). This toolbox is a Bayesian extension of the original metacognitive efficiency model (21) and provides robust parameter estimates in the face of uncertainty inherent in clinical groups of small sample size and relative heterogeneity (18).

### Perceptual decision-making

We also complimented the analysis of perceptual (first-order cognitive) decisions by estimating latent components of the decision-making process using the hierarchical drift diffusion model (HDDM) (22). Like HMeta-d, HDDM is particularly well suited to clinical research studies because it captures sources of uncertainty in the data (e.g., small group size and heterogeneous group features) in the form of posterior probability distributions of the parameter estimates. HDDM uses the choice and reaction time data to calculate latent parameters which estimate *how* individuals made perceptual decisions during the task. This was implemented in the openly available HDDM python toolbox (v0.8.0). Full details of the implementation process, model comparison and validation, and results of this approach are available in Supplementary Information.

### Statistical power

We powered this study *apriori* to detect a difference in metacognitive insight based on the effect size obtained by Fleming et al. (2014) as there are no published findings in HD. Their study detected differences across two clinical groups and controls using the same task and analysis method. We estimated the effect size (Cohen’s *f =* 0.53, α = 0.05, two-tailed) based on reported means (23). This revealed that a total sample size of 39 was required to achieve power of 0.8.

## RESULTS

### Participant demographics

Five participants were excluded prior to the analysis; four early-manifest HD patients were excluded due to saccadic impairment and one individual with premanifest HD was excluded due to a technical error while they completed the task. Included participants (N=58) were well-matched for age and sex across the groups (Table 1). All participants had MMSE scores in the normal range, but the early-manifest HD group had lower scores (*H*(2)=10.5, *p*=0.005). Premorbid verbal IQ was significantly lower in the pre- and early-manifest groups (*F*(2, 54)=5.2, *p*=0.009). Linear regression models were later used to understand if these differences were related to metacognitive efficiency. The early-manifest group had lower total functional capacity (TFC) scores than premanifest HD patients, as expected (W=164, p<0.01). Three of the early manifest patients and one premanifest gene-carrier were taking low-dose Olanzapine (2.5-5mg/day) for clinical reasons relating to their condition.

**Table 1.** Participant demographics. Groups were matched for age and sex. Groups had clinically normal, yet statistically different general cognitive and verbal IQ scores. The premanifest and early-manifest patients were different in their total UHDRS motor scores and functional capacity, as expected. Bolded p-values indicate significance at p<0.05. *Abbreviations: MMSE = Mini-Mental State Examination; UHDRS = Unified Huntington’s Disease Rating Scale; TFC = Total Functional Capacity. *One premanifest individual had an unusually high motor score (12) due to an unrelated hand injury. **One premanifest individual did not complete the National Adult Reading Test for verbal IQ so this cell contains one fewer measurement.*

|  | Premanifest<br>HD (N=19) | Early-manifest HD<br>(N=10) | Control<br>(N=29) | Test<br>Statistic | p-value |
| --- | --- | --- | --- | --- | --- |
| Age |  |  |  | 1.5 | 0.226 (1) |
| - Mean | 47.8 | 55.9 | 51.6 |  |  |
| - Range | 28.7 - 75.4 | 37.2 - 67.0 | 29.3 - 73.4 |  |  |
| Sex, Female | 11 (57.9%) | 7 (70.0%) | 12 (41.4%) | 2.9 | 0.238 (2) |
| MMSE |  |  |  | 10.5 | **0.005 (3)** |
| - Mean | 29.7 | 28.6 | 29.7 |  |  |
| - Range | 28.0 - 30.0 | 26.0 - 30.0 | 28.0 - 30.0 |  |  |
| Premorbid Verbal IQ |  |  |  | 5.2 | **0.009 (1)** |
| - Mean | 113.5** | 111.6 | 118.1 |  |  |
| - Range | 100.0 - 127.0 | 104.0 - 124.0 | 107.0 - 127.0 |  |  |
| UHDRS Total Motor |  |  |  | 4 | **<0.001(4)** |
| - Mean | 2.3 | 14.7 | - |  |  |
| - Range | 0.0 - 12.0* | 6.0 - 26.0 | - |  |  |
| TFC |  |  |  | 164 | **<0.001(4)** |
| - Mean | 12.8 | 11.4 | - |  |  |
| - Range | 11.0 - 13.0 | 10.0 - 13.0 | - |  |  |

1. Linear Model ANOVA 2. Pearson's Chi-squared test 3. Kruskal-Wallis one way ANOVA 4. Wilcoxon Mann-Whitney Rank Sum test

### Behavioural analysis

To assess behavioural performance, we compared mean accuracy (% correct), stimulus strength (Δ dots), response time and confidence ratings using one-way ANOVA or Kruskal-Wallis tests as non-parametric equivalent (see Supplementary Information for methods of statistical test selection). The staircase procedure successfully matched accuracy (% correct; Figure 2A) across the groups *(H(*2, 55) = 1.91, *p* = 0.38, η^2^ = 0.06). However, the mean stimulus strength to achieve that performance differed significantly between the groups (*F*(2, 55) = 13.85, *p* < 0.001, η^2^ = 0.33; Figure 2B). Pairwise comparison with Bonferroni correction method showed that patients with early-manifest HD (mean = 7.13 ± SEM = 0.4) completed the task with significantly greater stimulus strength (i.e., reduced difficulty level) compared with the premanifest group (mean = 5.68 ± SEM = 0.29; 95% CIs of mean difference = 1.25 - 3.53, adjusted *p* < 0.001), and also compared with healthy controls (mean = 4.74 ± SEM = 0.23; 95% CIs of mean difference = 0.24 - 2.67, adjusted *p* = 0.014). Further, the premanifest group performed with a significantly greater stimulus strength than the control group (95% CIs of mean difference = 0.02 - 1.86, adjusted *p =* 0.043). This shows that individuals with premanifest and early-manifest HD were impaired in making perceptual decisions compared to healthy controls. There were no significant differences in mean response time (*F*(2, 55) = 2.03, p=0.14, η^2^ = 0.07; Figure 2C). However, the trend towards reduced response time with manifest HD was further explored using the HDDM. There were also no differences in confidence level across the groups (*F*(2, 55) = 0.34, *p* = 0.71, η^2^ = 0.01; Figure 2D). This confirms that all participants were able to execute the perceptual decision and use the confidence scale as instructed. In addition, task accuracy was also matched across the groups throughout the entire experiment. There were no differences in accuracy across the eight task blocks (*F*(7, 440) = 0.59, *p* = 0.77, η^2^_p_ = 0.01), and no interaction effect of group by block (*F*(14, 440) = 1.02, *p* = 0.43, η^2^_p_ = 0.03).

### Perceptual decision-making model

We compared a limited number of regression models in order to determine the best-fitting HDDM to perceptual reaction time data. The best fitting model (lowest BPIC and DIC; Supplementary Table 1) was characterised by a regression in which drift rate was modulated by group and stimulus strength, their interaction, and decision threshold was modulated by group. Model parameters were reproducible (Supplementary Table 2) and simulated reaction time data based on these also accurately reproduced response times observed in our participants, including the trend towards faster response times with manifest HD (Supplementary Figure 2). Analysis of the posterior distributions of model parameters showed that healthy controls responded to stronger evidence (Δ dots, z-scored within-subjects) by significantly increasing their rate of evidence accumulation (drift rate), compared to both premanifest (*P* < 0.001) and early-manifest gene-carriers (*P* < 0.001) who did not differ (*P* = 0.34). Furthermore, premanifest gene-carriers set significantly lower decision thresholds for evidence accumulation than controls (*P* < 0.001), an impairment which was significantly greater in those with early-manifest disease (*P* < 0.001, Supplementary Figure 3).

### Metacognitive insight

M-ratio for each group was estimated separately and a higher value indicated better metacognitive insight. To assess if meaningful differences existed between the groups, we calculated 95% high density intervals (HDI) of differences between two distributions in pair-wise comparisons and compared the resulting difference distribution with 0. If the 95% HDI excluded 0, we considered this to be a meaningful (significant) difference.

There was no difference in metacognitive efficiency (M-ratio) between healthy controls (M: 0.68) and premanifest HD gene-carriers (M: 0.82; *P* = 0.1, 95% HDIs: −0.095 - +0.388). There was also no difference between the early-manifest HD gene-carriers (M: 0.79) and the control group (*P* = 0.25, 95% HDIs: −0.282 - +0.475). M-ratio was not reduced with greater disease burden, since early-manifest HD gene-carriers did not significantly differ from the premanifest group (*P =* 0.59, 95% HDIs: −0.458 - +0.34; Figure 3).

**Figure 3.**
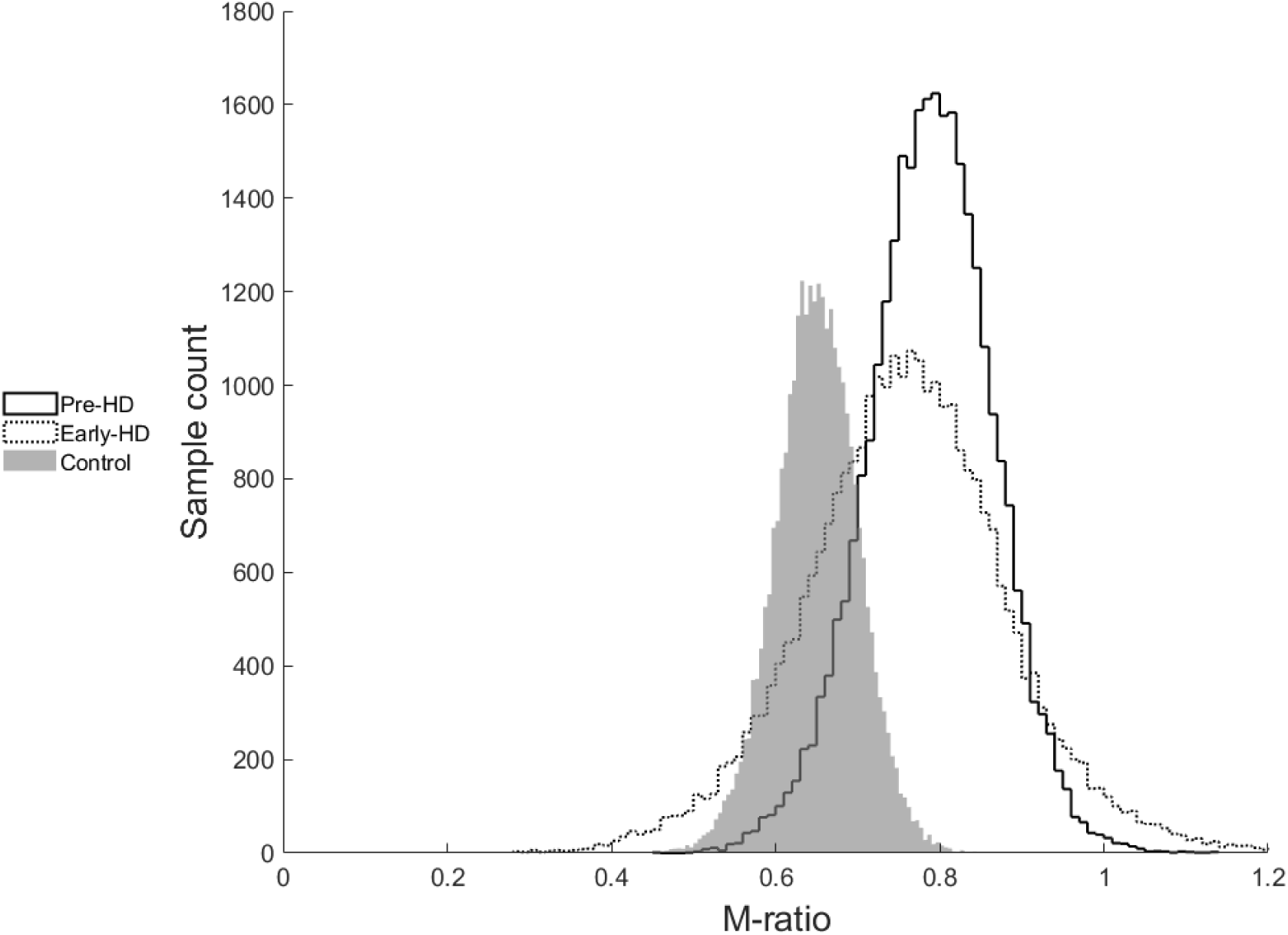
M-ratio sample estimates across the groups. There is significant overlap in the distributions indicating that gene-carriers showed similar metacognitive insight to controls.

To understand the contribution of individual differences, we conducted a post-hoc regression analysis in which metacognitive parameters (“M-ratio”, “metacognitive sensitivity”, “perceptual sensitivity”, “confidence”) were dependent variables. Predictors were HD gene status and several clinical covariates (age, gender, IQ, MMSE score, HADS-Anxiety score, HADS-Depression score). Continuous predictor variables were z-scored prior to the regression. Significance level for each regression model was adjusted using Bonferroni correction for the number of dependent variables (0.05/4 = 0.0125). This confirmed the previous finding that HD patients had intact metacognitive insight. A genetic diagnosis of HD was a significant predictor of improved metacognitive efficiency (*β* = +0.096, *p* = 0.007) after controlling for confounding individual differences *(R*^*2*^ = 0.43, *p* < 0.001; Figure 5). We found that HD gene status (*β* = +0.114, *p* = 0.003) was also a significant positive predictor of metacognitive sensitivity (*R*^*2*^ = 0.4, *p* < 0.001) but did not predict perceptual sensitivity (*R*^*2*^ = 0.06, *p* = 0.83). Since metacognitive efficiency is simply the ratio between metacognitive and perceptual sensitivity (M-ratio), this confirms that intact metacognitive efficiency in HD gene-carriers was driven by *increased metacognitive* sensitivity (*meta-d’*) and not *reduced perceptual* sensitivity (*d’*). Mean confidence was not directly associated with HD gene status, age, gender, IQ, cognition, anxiety or depression (*R*^*2*^ *=* 0.22, *p =* 0.09).

**Figure 4.**
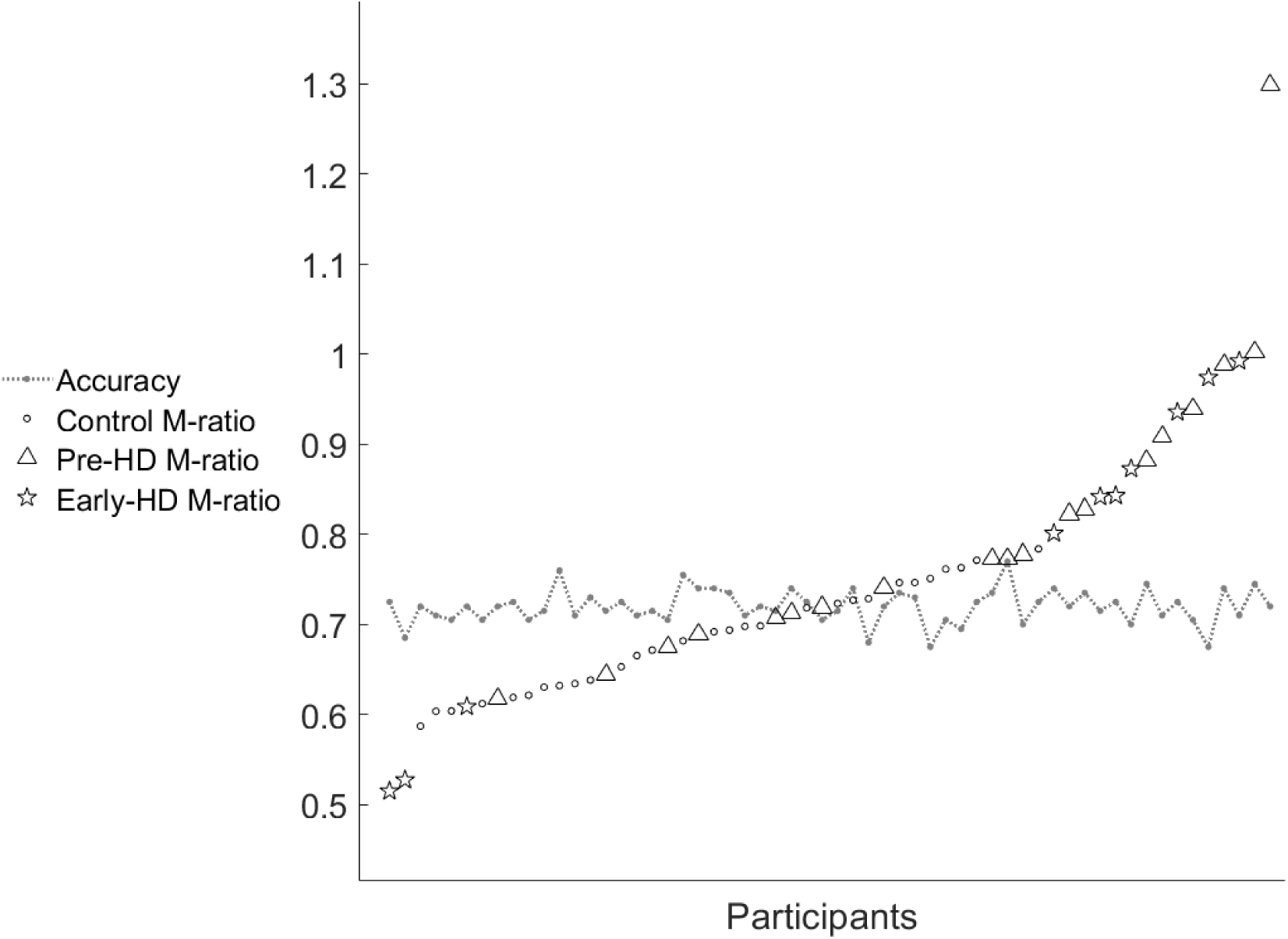
Individual mean accuracy (proportion correct) controlled at approximately 0.71 and mean M-ratio estimates. Each participant is a point on the X-axis.

**Figure 5.**
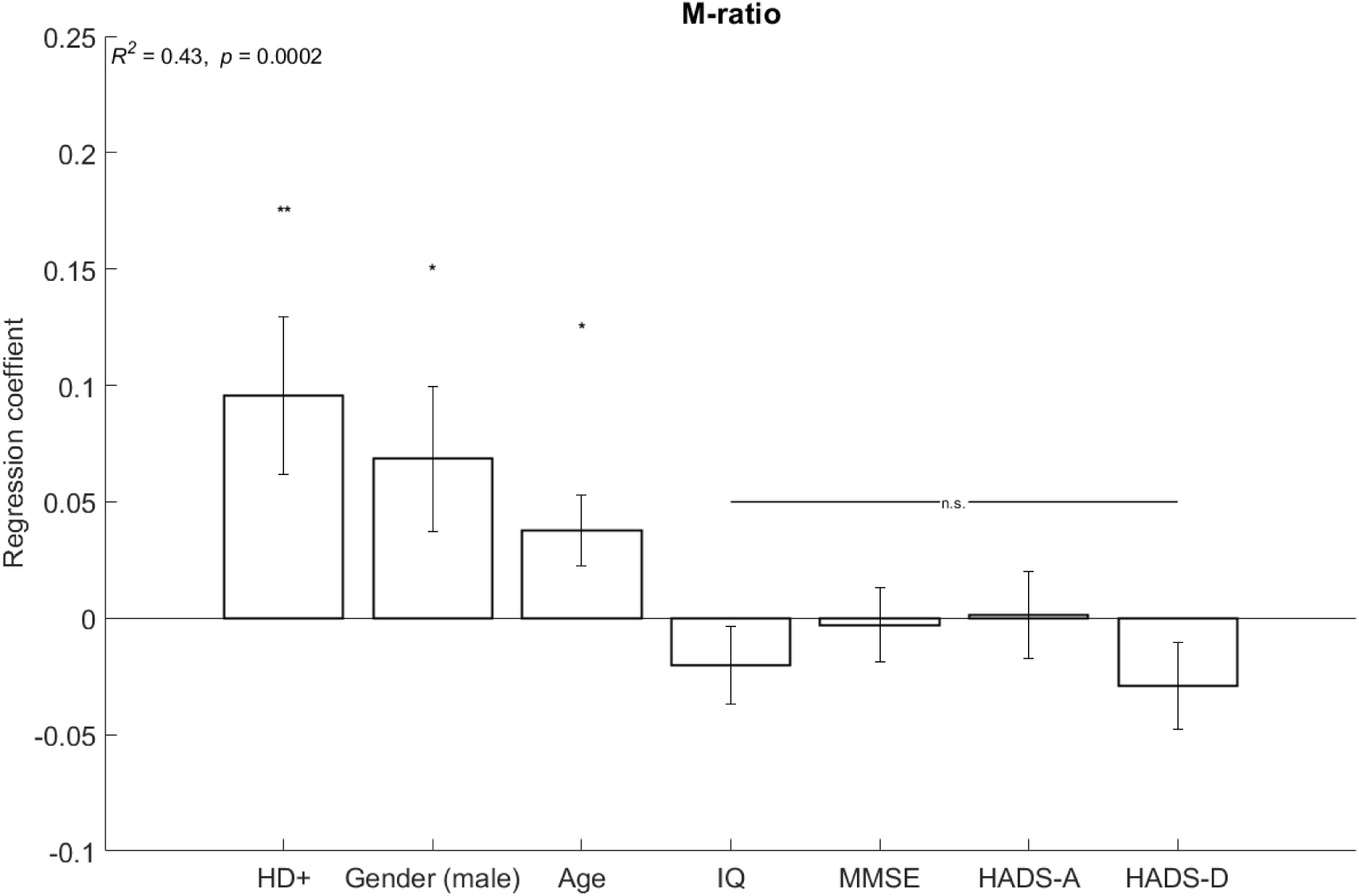
Linear regression coefficients for M-ratio (metacognitive efficiency) with independent predictors: HD gene status, age, gender, IQ, MMSE (Mini-Mental State Examination) score, HADS-Anxiety and HADS-Depression (Hospital Anxiety and Depression Score). n.s. = not significant, \**p* <0.05 and \*\**p* <0.01. Error bars indicate SEM.

## DISCUSSION

We report two novel findings about HD. Firstly, there is a deficit in perceptual decision-making that can be seen in the premanifest stage of the condition and gets worse in manifest disease, indicating that it is a product of the disease process rather than a genotype effect. Secondly, despite impaired perceptual decision-making performance, both premanifest and manifest HD gene-carriers demonstrated similar metacognitive insight into their performance compared to controls. In summary, we report a dissociation between impaired first-order cognition and intact, second-order, metacognition (trial-by-trial insight) in premanifest and early-HD gene-carriers.

HD gene-carriers required the perceptual decisions to be made objectively easier in order to perform as well as controls. Further, a computational model revealed this was underlined by impairments in evidence accumulation and reduced evidence thresholds. This was expected, as early manifest HD patients are impaired in the identification of ambiguous shapes and objects (24) and both premanifest and manifest gene-carriers show impairments in the recognition of faces and emotions (25–27).

In contrast, we predicted that metacognitive insight would be impaired in HD gene-carriers but found evidence to reject this hypothesis. Posterior distributions of metacognitive efficiency across all 3 groups did not differ. In a post-hoc analysis, having the HD gene was a significant predictor of improved metacognitive efficiency after controlling for the influence of age, gender, IQ, cognition, anxiety and depression. This was due to increased metacognitive sensitivity in HD gene-carriers and not reduced perceptual sensitivity. Age and gender were also significant predictors of metacognitive efficiency but IQ, cognition, anxiety and depression were not (Figure 5).

A possible explanation for intact metacognitive performance (despite impaired perceptual decision-making) is that a genetic diagnosis of HD induces a prior belief of current or future impairment and this leads to increased vigilance to performance-either consciously or subconsciously. In line with this, gene-carriers and their families often report “symptom hunting” and it is possible that trial-by-trial metacognitive insight in cognitively unimpaired individuals is attuned by this. However, we found no evidence of a negative confidence bias in gene-carriers (i.e., generally lower confidence; Figure 2D). Although intact metacognitive insight in HD gene-carriers was contrary to our hypothesis, other recent studies have identified performance improvements associated with HD gene-expansion. For example, Huntingtin gene expansion in low pathological ranges is associated with improved cognitive test scores and superior IQ performance in far-from-onset gene-carriers (28,29).

Intact metacognitive insight despite (impaired) cognitive performance in premanifest and early-HD is of clinical interest because it may be used to enhance subjective well-being and mental health (5). HD causes a wide range of psychological difficulties, but the literature on psychological interventions for people affected by HD is extremely limited (30). A recent feasibility study has shown that mindfulness-based cognitive therapy (which exploits metacognition) can be beneficial to individuals with premanifest HD (31). Our finding that early-HD gene-carriers retain good metacognitive insight further indicates that psychological therapies designed to apply this skill positively, may help maintain psychological well-being following a genetic diagnosis of HD.

## Limitations

The aim of this study was to assess whether local (trial-by-trial) metacognitive insight into cognitive performance is affected in the early stages of the HD disease process. We have shown that in relatively high functioning HD gene-carriers, metacognitive insight into cognitive performance is intact even though the performance itself is impaired. However, these findings relate only to HD gene-carriers who have *not* developed marked functional and cognitive impairments. Metacognitive insight may well decline as HD progresses. Consistent with this, there was increased uncertainty in the M-ratio for the early-manifest HD group; the posterior distribution is wider, with longer tails (Figure 3). This is likely due to the smaller sample size and greater heterogeneity of this group.

Secondly, changes in metacognitive performance may still occur early in HD in other cognitive domains or over different timescales (e.g., global insight). Research into metamemory in Alzheimer’s dementia has shown that local (i.e., trial-by-trial) metacognitive estimates are intact but global self-estimates are altered (32). Future studies should consider the progression between (early-stage, intact) local and (later-stage, impaired) global metacognitive insight in HD gene-carriers.

We did not include medication effects in our analyses. Dopamine is well-known to affect cognition (33) and manifest HD patients are often prescribed dopamine antagonists to help with the disease features, but these can increase the rate of cognitive decline (34). However, only 4 of 29 (13.8%) gene-carriers in this study were taking anti-dopaminergic medication, and all at low dose, so the pattern of findings cannot be explained by this.

## Conclusion

By dissociating perception and metacognition in HD, we show that perceptual decision-making impairments exist in HD gene-carriers without any other obvious symptoms or signs. However, metacognitive insight into cognitive performance remains intact, even in those who have progressed to manifest disease. Low-level perceptual issues which appear early in the disease may drive higher-order cognitive deficits that are often seen in the HD clinic. However, since metacognition is closely related to well-being and quality of life, clinicians and researchers should investigate how to exploit the high degree of local metacognitive insight that early HD gene-carriers can demonstrate.

## Data Availability

The task and HMeta-d toolbox for implementation in Matlab can be accessed thanks to Meta-Lab, UCL (https://github.com/metacoglab). Data and scripts used for data analysis in this study are available (https://github.com/samrchewitt/HD_perception_metacognition). Tutorials and installation guides for the HDDM toolbox (http://ski.clps.brown.edu/hddm_docs/). We encourage readers to contact the corresponding authors if they would like advice on applying or replicating this approach.

https://github.com/metacoglab/HMeta-d

https://github.com/metacoglab/meta_dots

http://ski.clps.brown.edu/hddm_docs/

## Acknowledgements

We would like thank the staff and study participants at the Huntington’s disease clinic at the John Van Geest Centre for Brain Repair who gave their time for this study.

## Data availability statement

the task and HMeta-d toolbox for implementation in Matlab can be accessed thanks to Meta-Lab, UCL (https://github.com/metacoglab). Data and scripts used for data analysis in this study are available (https://github.com/samrchewitt/HD_perception_metacognition). Tutorials and installation guides for the *HDDM* toolbox (http://ski.clps.brown.edu/hddm_docs/). We encourage readers to contact the corresponding authors if they would like advice on applying or replicating this approach.

