## Supplementary Information for "Metacognitive insight into cognitive performance in pre and early-stage Huntington’s disease"

#### Perceptual decision-making statistical power calculation

In addition to an *apriori* power calculation to detect differences in metacognitive insight between the groups, we confirmed that this study would also be powered to detect differences in perception based on O'Donnell and colleagues (4), who found deficits on a similar two alternative forced choice task in 2 groups of HD gene carriers and controls. Since effect size was not reported, we used G-Power software 3.1.9 to estimate the effect size (Cohen's $f$ = 0.44, $\alpha$ = 0.05, two-tailed) based on reported means. This indicated that a total sample size of 54 was required to achieve power of 0.8.

#### Behavioural Analysis

To determine if ANOVA was appropriate, normality of the behavioural data was confirmed in Matlab using the package *normalitytest*. This implements 10 independent normality tests (Kolmogorov-Smirnov test (Limiting form-Stephen's method, Marsaglia method), Lilliefors test, Anderson-Darling test, Cramer-Von Mises test, Shapiro-Wilk test, Shapiro-Francia test, Jarque-Bera test, D'Agostino and Pearson test). Data from each group was separately tested for normality and considered to come from a normal distribution if zero tests indicated a significant deviation from normality. Homogeneity of variance was subsequently confirmed with Bartlett's test (1). All pairwise comparisons were adjusted with a Bonferroni correction method. Eta squared effect sizes ($\eta^2$) were calculated in MATLAB from the sum of squares (SS) values in the ANOVA table output with the formula:

$$\eta^2 = SS_{effect} / SS_{effect} + SS_{error}$$

#### Perceptual decision-making model

The Hierarchical Drift Diffusion model (HDDM) simulates two-alternative forced choices as a noisy process of evidence accumulation through time, where sensory information is presented and the participant determines whether this provides evidence for either choice (2). Group-level parameters are estimated based on behavioural data (response time and choice accuracy), under the assumption that participants within a group are similar, but not identical, to each other. Parameter estimates are therefore constrained by group-level distributions. The rate of evidence accumulation is determined by the drift rate (v) parameter. Higher drift rates are related to faster and more accurate choices. A choice is made once the evidence reaches a decision boundary (a), which indicates the information threshold required to execute a decision and is related to response caution, with higher threshold indicating slower, more accurate choices. A third parameter, bias (z) indicates a starting point likelihood towards one boundary. The final parameter estimated is non-decision time (t), which captures decision-independent processing time (Supplementary Figure 1). This analysis was implemented in the openly available HDDM python toolbox (v0.8.0).

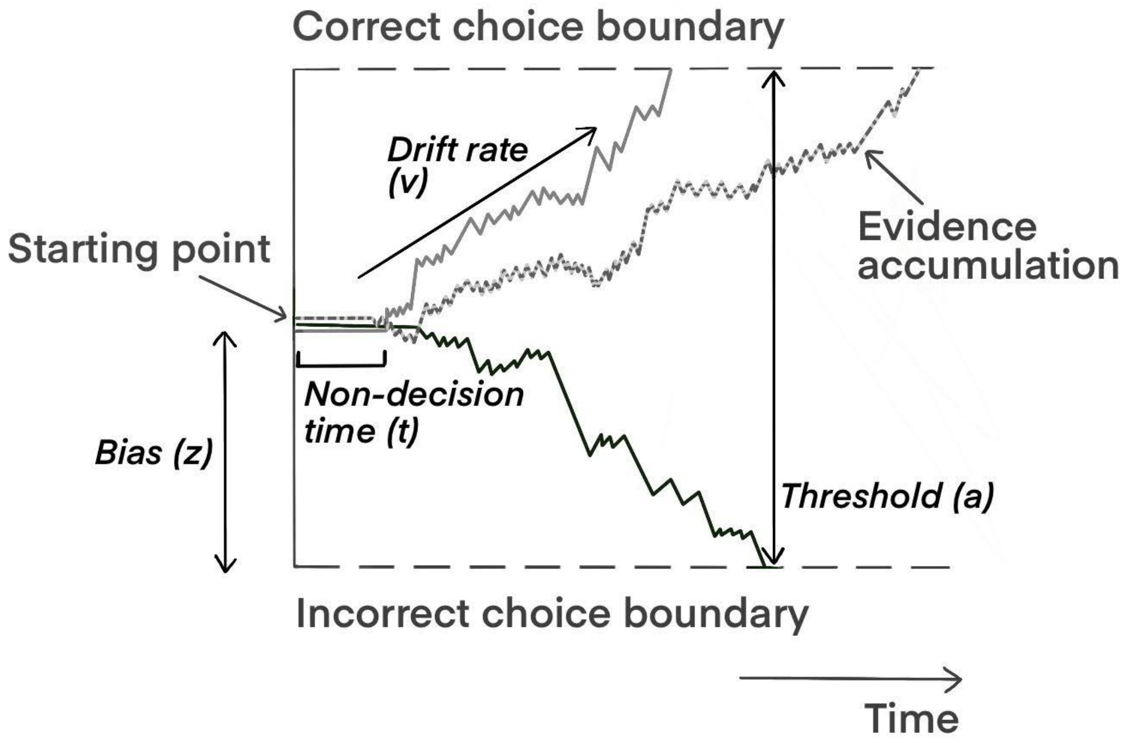


*Supplementary Figure 1. The hierarchical drift diffusion model was used to understand a decision between two choices as a noisy process of evidence accumulation through time. It calculates four latent parameters: drift rate (v; also called evidence accumulation), threshold (a), bias (z) and non-decision time (t). Information accumulates towards one of two boundaries (separated by a) with an average drift rate (v). Bias indicates the starting point likelihood towards one boundary. The flat line which precedes evidence accumulation (t) represents non-decision time, which includes time to encode stimuli and execute a motor response. This schematic shows three representative examples and not real data. Figure adapted from (3).*

Model comparison and validation

The best-fitting model to our data was determined by implementing several regression models within HDDM, in which responses were coded as correct and incorrect choices and drift rate (v) was modulated by stimulus strength on every trial (Supplementary Table 1). This is because we manipulated trial-by-trial stimulus strength and this is known to directly influence accumulation of evidence (5–7). The bias parameter was not included because by design, the task controlled the likelihood of a decision being correct or incorrect.

To test our hypothesis that HD gene carriers would show impairments in perceptual decision-making, we tested for a decoupling between evidence accumulation rate and the evidence presented to them. To do so, Z-scores of stimulus strength were calculated within subjects. Therefore, each participant had their own Z-scores, reflecting the distribution of evidence (stimulus strength) they were presented with across the experiment. This allowed us to determine the relationship between drift rate in individuals carrying the HD gene, without the confounding influence of absolute differences in stimulus strength, which we explicitly manipulated in order to control perceptual task performance (Δ dots; Figure 2B).

| Model | DIC | BPIC |
|---|---|---|
| v ~ stimulus strength | 26588.1 | 26592.1 |
| v ~ stimulus strength + group | 26571.2 | 26577.2 |
| v ~ stimulus strength + group + stimulus strength*group | 26573.3 | 26581.3 |
| **v ~ stimulus strength + group + stimulus strength*group, α ~ group** | **26392.9** | **26403** |
| v ~ stimulus strength + group + stimulus strength*group,<br>α ~ group,<br>*t* ~ group | 32486.2 | 32497.1 |

*Supplementary Table 1. DIC and BPIC values for each regression model implemented in HDDM. Values displayed are rounded to 1 decimal place. BPIC is calculated as (DIC + effective number of parameters (pD), and therefore provides a (2-fold) stricter penalty for additional complexity. The best fitting model (bold) included effects of stimulus strength and group on drift rate (v), and their interaction, plus an additional effect of group on decision threshold (a). Stimulus strength was Z-scored within participants.*

To address potential collinearity among parameters we fitted each model by estimating only group level posteriors for each regression coefficient, rather than for individual participants. Each regression model was sampled with 20,000 chains with the first 1000 chains discarded to estimate each parameter distribution. We defined the best-fitting model as that with the lowest DIC and BPIC (*bold text,* Supplementary Table 1). This model was characterised by a regression in which drift rate was modulated by group and stimulus strength, their interaction, and decision threshold was modulated by group:

$$v \sim Z_{\text{stimulus strength}} + \text{group} + (Z_{\text{stimulus strength}} * \text{group})$$

$$\alpha \sim \text{group}$$

Prior to analysing the posterior distributions of the best fitting model, we confirmed the model's reproducibility. We ran four, independent models in parallel to confirm the convergence of the resulting parameters using Rhat statistic. Rhat (or Gelman-Rubin) statistic is the ratio of the variance of each parameter when pooled together across the four models, to the within model variance. Therefore, Rhat quantifies the extent to which separate models reach different conclusions (8). Model parameters demonstrated excellent convergence for all estimated parameters (mean: 1.00003, range: 0.99998 - 1.00015; Supplementary Table 2). Satisfied with this, we combined the chains of the four models and analysed the posterior distributions of the combined best-fitting model, which increased the sample size for the parameter estimates (80,000 chains, initial 4000 discarded). Of note, a model with a group term for non-decision time was a poorer fit to our data, which suggests that non-decision time did not differ between the groups.

| | **Model 1** | | **Model 2** | | **Model 3** | | **Model 4** | | **Combined** | | |
|---|---|---|---|---|---|---|---|---|---|---|---|
| | mean | std | mean | std | mean | std | mean | std | mean | std | Rhat |
| **Non-decision time (t)** | 0.499 | 0.003 | 0.498 | 0.003 | 0.498 | 0.003 | 0.499 | 0.003 | 0.498 | 0.003 | 1.0002 |
| **v Control** | 0.562 | 0.016 | 0.561 | 0.016 | 0.561 | 0.016 | 0.562 | 0.016 | 0.562 | 0.016 | 1.0000 |
| **v Pre-HD** | 0.614 | 0.021 | 0.614 | 0.021 | 0.614 | 0.021 | 0.614 | 0.021 | 0.614 | 0.021 | 1.0000 |
| **v Early-HD** | 0.595 | 0.030 | 0.595 | 0.030 | 0.596 | 0.030 | 0.595 | 0.030 | 0.595 | 0.030 | 1.0000 |
| **v Control*stimulus** | 0.243 | 0.015 | 0.243 | 0.016 | 0.243 | 0.016 | 0.243 | 0.016 | 0.243 | 0.016 | 1.0000 |
| **v Pre-HD*stimulus** | -0.015 | 0.026 | -0.015 | 0.026 | -0.015 | 0.026 | -0.015 | 0.026 | -0.015 | 0.026 | 1.0000 |
| **v Early-HD*stimulus** | 0.000 | 0.033 | 0.001 | 0.034 | 0.000 | 0.034 | 0.000 | 0.034 | 0.000 | 0.034 | 1.0000 |
| **threshold (a) Control** | 1.990 | 0.014 | 1.991 | 0.014 | 1.991 | 0.014 | 1.990 | 0.014 | 1.991 | 0.014 | 1.0000 |
| **threshold (a) Pre-HD** | 1.887 | 0.016 | 1.887 | 0.015 | 1.888 | 0.016 | 1.887 | 0.016 | 1.887 | 0.016 | 1.0000 |
| **threshold (a) Early-HD** | 1.690 | 0.018 | 1.690 | 0.018 | 1.690 | 0.018 | 1.690 | 0.018 | 1.690 | 0.018 | 1.0000 |

*Supplementary Table 2. Mean, standard deviation and Gelman Rubin statistic (Rhat) of the HDDM parameters from four best-fitting models estimated independently. Rhat Values < 1.1 are considered to indicate acceptable convergence (8). Rhat statistics indicate that the model parameters are highly reproducible. v=drift rate.*

A further validation of the model is that, based on the parameters, we are able to reproduce the behaviour of our participants. To confirm this, we performed a posterior predictive check in which we simulated response time distributions generated from the posterior distributions of the model parameters and compared them with observed response times. HDDM simulates 500 response time distributions for each participant independently and quantiles are the mean across all simulations. Taking all participants together, the model reproduced response times accurately. This was also the case for each group, and for both correct and incorrect responses (Supplementary Figure 2). For example, the model reproduces the (non-significant) trend toward faster response times with HD (See main text, Figure 2).

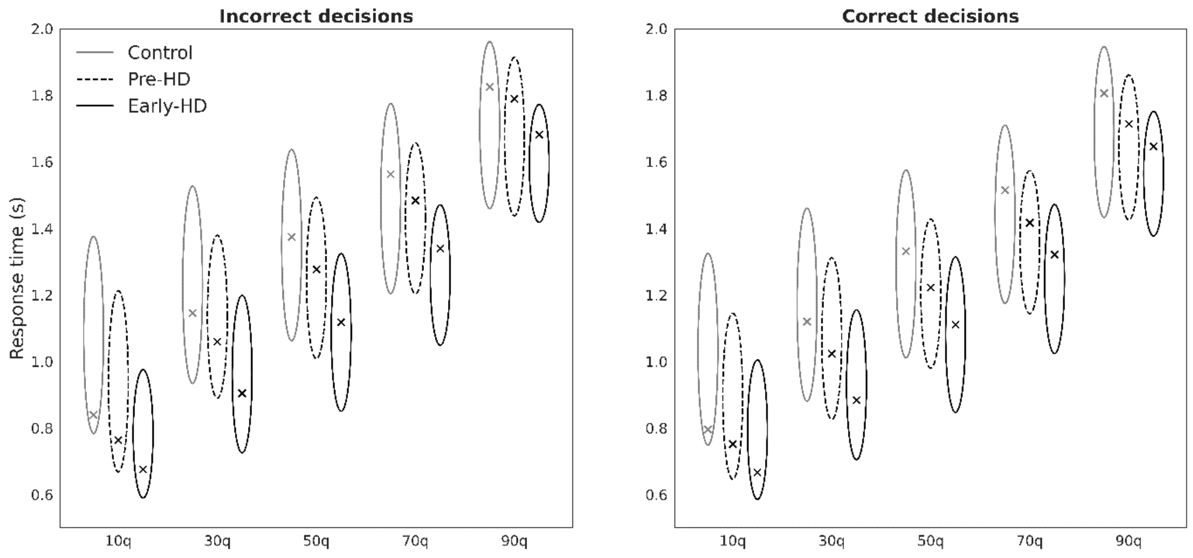


*Supplementary Figure 2. Simulated response times reproduced the empirically observed response times across all groups and the entire distribution of responses. Group mean response times for each quantile are plotted as X and simulated response times from the posterior predictive of the HDDM as ellipses (capturing uncertainty). All observed means fall within the model's posterior predictions. Quantiles are computed for each subject separately and averaged to yield group quantiles. Ellipse lengths are determined by the standard deviation of the posterior predictive distribution for that quantile and group. All ellipse widths are equal (0.1 SD).*

Posterior distribution analysis

To assess if meaningful differences in parameter estimates existed between the groups, we compared the posterior distributions of each group directly and calculated the probability that the difference between the group distributions was in the opposite direction. This is similar to a one-tailed t-test (we calculated the probability, *P,* that the distribution with the greater mean was in fact smaller) and considered probability (*P*) < 0.025 (one-tailed) as statistically significant.

At the group level (considering all trials equally), there was a significant increase in the drift rate parameter in the premanifest group (M = 0.614, SD = 0.021) compared with the control group (M = 0.561, SD = 0.016; *P* = 0.022). Drift rate in the early-manifest group did not significantly differ from the control group (M = 0.595, SD = 0.03, *P* = 0.16) or the premanifest group (*P* = 0.31; Supplementary Figure 3A).However, such overall group differences do not take into account differences in stimulus strength (Δ dots) between the groups (Supplementary Figure 3B), which we explicitly manipulated. Consistent with our hypothesis, we found that the interaction effect of group*$Z_{\text{stimulus strength}}$ on drift rate revealed significant differences between both HD groups and the controls. In controls (M = 0.243, SD = 0.016), the effect of increasing $Z_{\text{stimulus strength}}$ on drift rate was significantly greater than in the premanifest group (M = -0.015, SD = 0.026; *P* < 0.001), and the early-manifest group (M = 0, SD = 0.034; *P* < 0.001). In other words, compared with both HD groups, healthy controls responded to relatively stronger evidence in favour of the correct decision by accumulating evidence more quickly. There was no difference between the premanifest and the early-manifest groups (*P* = 0.34; Supplementary Figure 3B), implying that this deficit emerges early in HD and is stable between disease stages.

Comparing the decision threshold parameter, we found further significant differences between the groups. Patients with early-manifest HD adopted the lowest threshold (M = 1.69, SD = 0.018), which was significantly reduced compared to the premanifest gene-carriers (M = 1.89, SD = 0.016, $P < 0.001$) and the control group (M = 1.99, SD = 0.014, $P < 0.001$). The threshold adopted by the premanifest group was also significantly reduced compared to the control group ($P < 0.001$; Supplementary Figure 3C). In summary, decision thresholds were consistently narrowed with increased disease status.

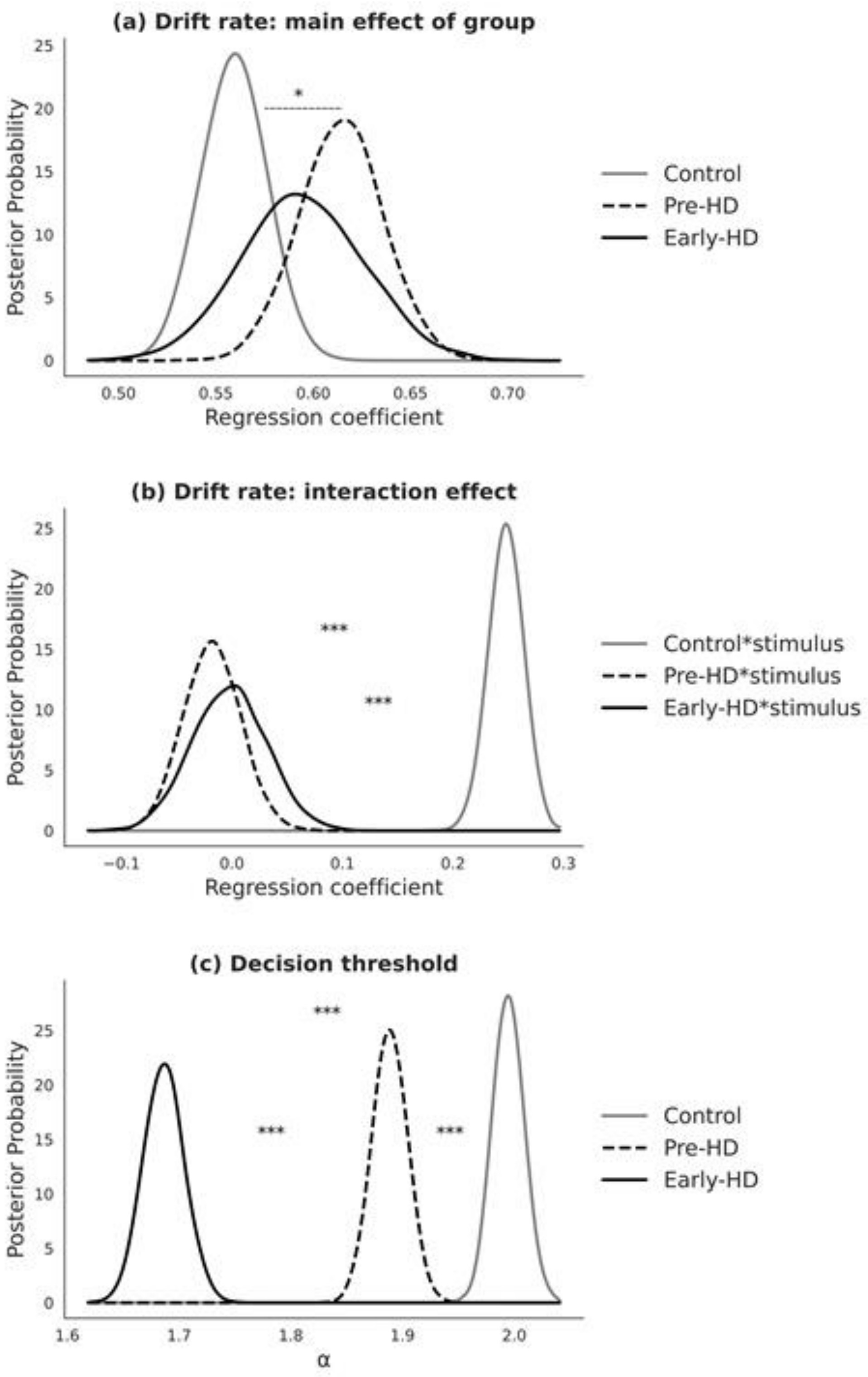


*Supplementary Figure 3. Posterior probability distributions from the best-fitting HDDM regression model. (a) Group level drift rates. (b) Significant interaction between drift rate and stimulus strength: the effect of increasing $Z_{stimulus\ strength}$ on drift rate in both premanifest HD and early-manifest HD was significantly reduced compared with the control group. (c) Significant reductions in decision threshold with greater disease status (c). *P(one-tailed) < 0.025. ***P(one-tailed) < 0.001.*

**Metacognition model**

One premanifest-HD participant had a high M-ratio which greatly exceeded the group mean (See main text, Figure 3). To confirm the effect that this participant had on the group estimate and therefore, our conclusions, we ran the HMeta-d analysis again with this participant excluded. The results were qualitatively and statistically equivalent. There was no difference between the posterior distributions derived from all premanifest HD participants (Main text, figure 3) and from the sample excluding this participant ($P$ = 0.59, 95% HDI: -0.28 - +0.22). There also remained no significant differences in M-ratio between healthy controls and the reduced sample premanifest-HD ($P$ = 0.118, 95% HDI: -0.09 - +0.34) or between the reduced sample premanifest HD and early-manifest HD ($P$ = 0.25, 95% HDI: -0.42 - +0.35). In other words, including this participant did not alter our conclusion that metacognitive insight into cognitive performance was intact in premanifest-HD.

### Supplementary Information References


1. Snedecor GW, Cochran WG. *Statistical Methods*. Eighth. Iowa State University Press (1989).
